# Correlations Between Community-Level HIV Preexposure Prophylaxis Coverage and Individual-Level Sexual Behaviors among US Men Who Have Sex with Men

**DOI:** 10.1101/2021.07.16.21260658

**Authors:** Laura M. Mann, Adrien Le Guillou, Steven M. Goodreau, Julia L. Marcus, Travis Sanchez, Kevin M. Weiss, Samuel M. Jenness

## Abstract

**Background:** HIV preexposure prophylaxis (PrEP) has been associated with changes in sexual behavior after PrEP initiation. However, behavioral differences may also emerge among PrEP non-users in communities with high PrEP coverage.

**Methods:** We used demographic, behavioral, and sexual network data from ARTnet, a cross-sectional study of US men who have sex with men conducted during 2017–2019. Multivariable regression models with a Bayesian modeling framework in which individuals were nested within their residential geographic areas were used to estimate associations between area-level PrEP coverage and five sexual behavior outcomes (number of total, main, and casual male partners [network degree]; count of one-time partnerships; and consistent condom use in one-time partnerships), controlling for individual PrEP use.

**Results:** PrEP coverage ranged from 10.3% (Philadelphia) to 38.9% (San Francisco). Total degree was highest in Miami (1.35) and lowest in Denver (0.78), while the count of one-time partners was highest in San Francisco (11.7/year) and lowest in Detroit (1.5/year). Adjusting for individual PrEP use and demographics, community PrEP coverage was associated with higher total degree (adjusted incidence rate ratio [aIRR]=1.73; 95% CrI, 0.92–3.44), casual degree (aIRR=2.05; 95% CrI, 0.90–5.07), and count of one-time partnerships (aIRR=1.90; 95% CrI, 0.46–8.54). Without adjustment for individual PrEP use, these associations were amplified. There were weaker associations with main degree (aIRR=1.21; 95% CrI, 0.48–3.20) and consistent condom use in one-time partnerships (aIRR=1.68; 95% CrI, 0.86–3.35).

**Conclusions:** Most of the associations between community PrEP coverage and sexual behavior were explained by individual PrEP use. However, there were residual associations after controlling for individual PrEP use, suggesting that PrEP coverage may partially drive community-level changes in sexual behavior.

## INTRODUCTION

The prevention of human immunodeficiency virus (HIV) remains a major public health challenge both globally and in the United States. The rate of HIV diagnoses in the US remains persistently high, with nearly forty thousand reported cases in 2019.^1^ Men who have sex with men (MSM) are at increased risk for HIV: despite representing less than 5% of the US population, MSM account for nearly 70% of all diagnoses.^1^ Preexposure prophylaxis (PrEP) significantly reduces the risk of HIV acquisition,^2^ but, despite substantially increasing levels of PrEP use,^3^ its impact on population-level HIV incidence in the US has been relatively modest.^4,5^ Empirical effects are much weaker than projections by mathematical modeling studies estimating incidence reduction associated with various PrEP coverage levels.^6^

Gaps between projected and actual HIV rates may be attributable to inequities in PrEP coverage,^3^ but they may also partially be driven by changes in sexual behaviors after PrEP initiation, often referred to as behavioral “risk compensation.”^7,8^ An individual, for example, may engage in more condomless sex after initiating PrEP because PrEP reduces their perceived (and actual) risk of acquiring HIV.^7^ A recent systematic review found broad evidence for an increase in condomless sex after participants commence PrEP.^9^ While behavioral changes as a result of PrEP have few implications for HIV incidence among active, adherent PrEP users, they have the potential to increase HIV acquisition in cases of suboptimal adherence or unplanned PrEP discontinuation.^10^ Further, behavioral changes increase the risk of acquiring other non-HIV sexually transmitted infections (STIs).^9^

Other PrEP-related changes in behavior may also contribute to the modest population-level impact of PrEP. With the continually increasing scale-up of PrEP, changes in risk perceptions and behavior may also occur among individuals not using PrEP but living in cities with high PrEP coverage. This phenomenon, which has been referred to as “community-level risk compensation,”^7^ could occur when individuals who are not on PrEP feel that they are less likely to acquire HIV if others in their community are protected against HIV and modify their behavior as a result. Resulting behavioral modifications could include changes to the number of sexual partners, the frequency of partnership formation, or the practice of condom use.

Behavioral changes among PrEP non-users may be akin to unvaccinated individuals feeling indirectly protected from vaccine-preventable diseases if others in their community are protected.^11^ These behavioral changes could have a considerable impact on population-level HIV incidence because PrEP coverage is very unequally distributed, even in communities where total PrEP coverage is high.^3^

Few studies have directly quantified evidence for community-level changes in behavior associated with PrEP coverage. Previous studies have used participant-reported population-level condom use after wide-scale PrEP introduction as an indicator for community-level behavioral changes, but did not adequately distinguish behavioral changes among individuals who were directly versus indirectly protected by PrEP.^12,13^ One study examined trends in condomless sex among MSM not on PrEP during large-scale PrEP implementation projects in Australia (2016–March 2017), but only considered condomless sex, only one form of behavior change.^14^ Previous studies have not estimated PrEP-related community-level behavioral differences using data from multiple communities or multiple behavioral outcomes; this approach is necessary as PrEP coverage may prompt modifications to a range of sexual behaviors and patterns may vary by community.

In this study, we examined the associations between community-level PrEP use and five behavioral indicators to determine whether PrEP-related community-level behavioral differences exist independent of individual-level PrEP use. Our study examines multiple sexual behavior outcomes, including those related to sexual networks. We hypothesized that communities with high PrEP coverage would have higher sexual network connectivity, lower condom use, and more one-time partners, even after adjusting for individual-level PrEP use. Our broader goal of this study was to elucidate the indirect effects of PrEP on sexual behaviors among US MSM in order to guide effective HIV and STI prevention efforts.

## METHODS

### Study Design

This study used data from ARTnet, a cross-sectional web-based US study of MSM conducted during 2017–2019.^15^ Participants were recruited through the American Men’s Internet Survey. ARTnet eligibility criteria included male sex at birth, current male identity, lifetime history of sexual activity with another man, and age 15–65 years. The study collected data on demographic and clinical characteristics, sexual behaviors, and egocentric network structures.^15^ The Emory University Institutional Review Board approved the study.

### Measures

Participants were asked summary questions about their overall number of male partnerships within three types in the past year: main (a “boyfriend, significant other, or life partner”), casual (a non-main partner they have had sex with more than once), and one-time. They were then asked detailed partner-specific questions for up to their five most recent partners. These questions included attributes of the partner (e.g., demographics) and the partnership itself (e.g., start/end dates, frequency of sexual activity).

From these partnership data, we calculated total, main, and casual network degree. Degree was the number of extant persistent partners (with receptive or insertive anal intercourse and/or oral sex), for each partnership type, on the day of the survey. We calculated one-time partnership acquisition by subtracting the reported main and casual partners from the total past-year partners. We also evaluated consistent condom use (always using condoms) in one-time partnerships. Individual PrEP information was based on self-reported current use (“Are you currently taking PrEP?”).

Our analysis was restricted to MSM who had ever had an HIV test and who self-reported as HIV-negative. Based on ZIP code of residence and by matching against county databases, individuals were classified as residing in one of 15 metropolitan statistical areas (major cities) or nine US census divisions. Cities and census division were mutually exclusive (e.g., New England contained all of New England but excluded the Boston MSA). This choice was made because community-level factors of interest may differ within versus outside of urban centers. Community PrEP coverage was calculated for each area; this represents the cross-sectional proportion of HIV-negative MSM in each geographic area that were currently on PrEP. We compared the estimates of community PrEP coverage to National HIV Behavioral Surveillance System (NHBS) estimates in the Supplemental Appendix to validate their accuracy.

### Statistical Analyses

All analyses were conducted using R 4.0.3. Descriptive demographic and behavioral variables were stratified by geography. We used a hierarchical Bayesian modeling framework in which individuals were nested within their geographic areas. Weakly informative prior distributions were placed on all model parameters. Models were fit with the *rethinking* package, which uses the STAN Markov Chain Monte Carlo Sampler to estimate model coefficients. Within the Bayesian models, we jointly estimated the outcomes of interest and community PrEP coverage with a binomial model of individual PrEP. This multilevel design by community accounted for the sample differences in the number of respondents by community. Our model outcomes were total degree, main degree, casual degree, count of one-time partners, and consistent condom use in one-time partnerships.

We used a multi-step approach to estimate individual-level versus community-level PrEP-related differences in sexual behaviors. First, to model individual-level PrEP-related sexual behavior differences, we used multivariable regression to estimate incidence rate ratios (IRRs) between individual-level PrEP use and the outcomes of interest, adjusting for potential demographic confounders (age and race). IRRs represent the ratio of the rate of the outcomes of interest (degree, partners, or consistent condom use, for the past year) comparing the indicated exposure scenarios. Second, to estimate community-level PrEP-related differences in behaviors, we examined PrEP coverage and behavior outcomes by city/region and used multivariable regression to estimate IRRs for the association between community PrEP coverage and our five outcomes of interest. We ran these models first adjusting for demographics only, then for demographics plus individual PrEP status. This strategy allowed us to evaluate the magnitude of the impact of individual-level PrEP status on the associations between community PrEP coverage and behavior. For models of total, main, and casual degree, and consistent condom use in one-time partnerships, Poisson regression was used. Due to overdispersion, negative binomial regression was used for the count of one-time partners; prevalence ratios that approximate IRRs were generated. Our analysis code is available in a git repository at https://github.com/EpiModel/PrEPCommunityLevelBehaviors.

Recent developments in epidemiology suggest that p-values often damage the interpretation of epidemiologic data and should not be used to evaluate associations.^16^ Therefore, we did not present p-values nor only deem 95% confidence intervals that did not contain 1.0 as epidemiologically meaningful. Instead, we used a Bayesian approach, reporting 95% credible intervals and the proportion of posterior distributions from simulations that indicate an association above 1.00 for each model. We interpreted results as signaling a meaningful pattern if the majority (approximately ≥80%) of the posterior distributions from the simulations indicated a positive association.

## RESULTS

Of the 3,259 HIV-negative MSM in our study, 631 (19.4%) reported current PrEP use, 178 (5.5%) reported previous but not current PrEP use, and the remaining 2,450 (75.2%) reported never using PrEP (Table 1). Participants ranged in age from 15–65 years, with an average age of 37.3 years. Approximately 74.3% of participants were non-Hispanic white, 12.9% were Hispanic, 4.1% were non-Hispanic Black, and 8.7% were non-Hispanic other race. Participants were from across the US, with the most represented census divisions being the South Atlantic (21.5%) and the Pacific (17.2%) divisions. Approximately 92.4% of participants reported having health insurance (public or private).

**Table 1.** Characteristics of HIV-negative ARTnet Participants Stratified by PrEP Use

|  | Total* | Current PrEP Use | Non-Current PrEP Use | Never Used PrEP |
| --- | --- | --- | --- | --- |
| Total persons | 3,259 (100.0%) | 631 (19.4%) | 178 (5.5%) | 2,450 (75.2%) |
| Race/ethnicity |  |  |  |  |
| Black, non-Hispanic | 134 (4.1%) | 28 (4.4%) | 4 (2.2%) | 102 (4.2%) |
| Hispanic | 422 (12.9%) | 81 (12.8%) | 23 (12.9%) | 318 (13%) |
| Other, non-Hispanic | 283 (8.7%) | 53 (8.4%) | 11 (6.2%) | 219 (8.9%) |
| White, non-Hispanic | 2,420 (74.3%) | 469 (74.3%) | 140 (78.7%) | 1,811 (73.9%) |
| Age (yr) |  |  |  |  |
| 15–24 | 713 (21.9%) | 71 (11.3%) | 36 (5.7%) | 606 (24.7%) |
| 25–34 | 947 (29.1%) | 196 (31.1%) | 76 (42.7%) | 675 (27.6%) |
| 35–44 | 518 (15.9%) | 122 (19.3%) | 26 (14.6%) | 370 (16.3%) |
| 45–54 | 573 (17.6%) | 149 (23.6%) | 24 (13.5%) | 400 (16.3%) |
| 55–65 | 508 (15.6%) | 93 (14.7%) | 16 (9.0%) | 399 (16.3%) |
| Census division |  |  |  |  |
| New England | 165 (5.1%) | 31 (4.9%) | 16 (9%) | 118 (4.8%) |
| Middle Atlantic | 442 (13.6%) | 92 (14.6%) | 20 (11.2%) | 330 (13.5%) |
| South Atlantic | 701 (21.5%) | 119 (18.9%) | 35 (19.7%) | 547 (22.3%) |
| East South Central | 132 (4.1%) | 19 (3%) | 6 (3.4%) | 107 (4.4%) |
| West South Central | 323 (9.9%) | 61 (9.7%) | 11 (6.2%) | 251 (10.2%) |
| East North Central | 466 (14.3%) | 92 (14.6%) | 25 (14%) | 349 (14.2%) |
| West North Central | 187 (5.7%) | 32 (5.1%) | 13 (7.3%) | 142 (5.8%) |
| Mountain | 281 (8.6%) | 30 (4.8%) | 21 (11.8%) | 230 (9.4%) |
| Pacific | 562 (17.2%) | 155 (24.6%) | 31 (17.4%) | 376 (15.3%) |
| Health insurance |  |  |  |  |
| Private | 2,441 (76.4%) | 516 (82.2%) | 128 (71.9%) | 1,797 (75.2%) |
| Public | 514 (16.1%) | 94 (15%) | 33 (18.5%) | 387 (16.2%) |
| None | 242 (7.6%) | 18 (2.9%) | 17 (9.6%) | 207 (8.7%) |
| Average total degree* | 1.04 (SD 1.07) | 1.58 (SD 1.28) | 0.98 (SD 0.88) | 0.91 (SD 0.97) |
| Median total degree | 1.00 | 1.00 | 1.00 | 1.00 |
| Average count of one-time partnerships | 4.73 (SD 12.1) | 12.8 (SD 21.3) | 5.62 (SD 11.9) | 2.57 (SD 6.7) |
| Median count of one-time partnerships | 1.00 | 5.00 | 2.00 | 0.00 |
| Screened for an STI in past 12 months |  |  |  |  |
| Yes | 1,677 (54.4%) | 547 (89.1%) | 122 (70.9%) | 1,008 (43.8%) |
| No | 1,408 (45.6%) | 67 (10.9%) | 50 (28.1%) | 1,291 (56.2%) |
| Screened for HIV in past 12 months |  |  |  |  |
| Yes | 2,106 (71.9%) | 561 (98.2%) | 129 (79.6%) | 1,416 (64.5%) |
| No | 822 (28.1%) | 10 (1.8%) | 33 (20.4%) | 779 (35.5%) |

PrEP, pre-exposure prophylaxis; SD, standard deviation; STI, sexually transmitted infection
\*Total degree is the number of persistent male partners, including main and casual partners, over the past year, measured on the day of the survey completion.

PrEP use varied by region/city, as demonstrated in Table 2. PrEP-eligible men living in San Francisco and Seattle had the highest current PrEP use (38.9% and 33.8%, respectively) and men in Philadelphia and Detroit had the lowest prevalence of current PrEP use (10.3% and 10.9%).

**Table 2.** PrEP Usage by Region/City for HIV-negative ARTnet Participants

|  | **Total** | **Currently on PrEP** | **Not on PrEP (Includes Non-Current and Never Used PrEP)** |
| --- | --- | --- | --- |
| Total | 3,259 | 631 (19.4%) | 2,628 (80.6%) |
| Census division/city<sup>†</sup> |  |  |  |
| Northeast |  |  |  |
| New England | 87 | 14 (16.1%) | 73 (83.9%) |
| Boston | 78 | 17 (21.8%) | 61 (78.2%) |
| Middle Atlantic | 159 | 28 (17.6%) | 131 (82.4%) |
| New York City | 211 | 56 (26.5%) | 155 (73.5%) |
| Philadelphia | 78 | 8 (10.3%) | 70 (89.7%) |
| South |  |  |  |
| South Atlantic | 371 | 45 (12.1%) | 326 (87.9%) |
| Atlanta | 136 | 32 (23.5%) | 104 (76.5%) |
| Miami | 55 | 7 (12.7%) | 48 (87.3%) |
| Washington, DC | 133 | 35 (26.3%) | 98 (73.7%) |
| East South Central | 132 | 19 (14.4%) | 113 (85.6%) |
| West South Central | 166 | 27 (16.3%) | 139 (83.7%) |
| Dallas | 84 | 17 (20.2%) | 67 (79.8%) |
| Houston | 73 | 17 (23.3%) | 56 (76.7%) |
| Midwest |  |  |  |
| East North Central | 284 | 47 (16.5%) | 237 (83.5%) |
| Chicago | 136 | 40 (29.4%) | 96 (70.6%) |
| Detroit | 46 | 5 (10.9%) | 41 (89.1%) |
| West North Central | 187 | 32 (17.1%) | 155 (82.9%) |
| West |  |  |  |
| Mountain | 226 | 24 (10.6%) | 202 (89.4%) |
| Denver | 55 | 6 (10.9%) | 49 (89.1%) |
| Pacific | 214 | 48 (22.4%) | 166 (77.6%) |
| Los Angeles | 132 | 36 (27.3%) | 96 (72.7%) |
| San Diego | 49 | 10 (20.4%) | 39 (79.6%) |
| San Francisco | 90 | 35 (38.9%) | 55 (61.1%) |
| Seattle | 77 | 26 (33.8%) | 51 (66.2%) |

PrEP: pre-exposure prophylaxis
<sup>†</sup>Regions include the nine US Census Bureau divisions but exclude the major listed cities within these divisions, where relevant. The New England division excludes Boston; the Middle Atlantic division excludes New York City and Philadelphia; the South Atlantic division excludes Atlanta, Miami, and Washington, DC; the East South Central division has no exclusions; the West South Central division excludes Dallas and Houston; the East North Central division excludes Chicago and Detroit; the West North Central division has no exclusions; the Mountain division excludes Denver; the Pacific division excludes Los Angeles, San Diego, San Francisco, and Seattle.

Behavioral differences were observed by region/city; summary measures of behavioral outcomes stratified by region/city are shown in Table 3. Additionally, behavioral outcomes varied by reported PrEP use. For example, men currently on PrEP had an average casual degree of 1.17 (SD 1.23), whereas men not on PrEP had an average casual degree of 0.48 (SD 0.90) (Table 3, Supplemental Table 1). Men currently on PrEP had a higher average count of one-time partnerships over the past year (12.8; SD 21.3) compared to men not on PrEP (2.8; SD 7.2).

**Table 3.** Sexual Network and Sexual Behavior Parameters for HIV-negative ARTnet Participants by Region/City

|  | Average Main Degree* | Average Casual Degree | Average Total Degree | Average Count of One-Time Partnerships | Proportion of Individuals Who Always Used Condoms in One-time Partnership(s) |
| --- | --- | --- | --- | --- | --- |
| Total | 0.43 | 0.61 | 1.04 | 4.73 | 0.44 |
| Census division/city† |  |  |  |  |  |
| Northeast |  |  |  |  |  |
| New England | 0.41 | 0.45 | 0.86 | 3.32 | 0.42 |
| Boston | 0.42 | 0.54 | 0.96 | 3.59 | 0.57 |
| Middle Atlantic | 0.44 | 0.59 | 1.03 | 4.83 | 0.51 |
| New York City | 0.38 | 0.67 | 1.06 | 5.35 | 0.54 |
| Philadelphia | 0.42 | 0.51 | 0.94 | 2.22 | 0.47 |
| South |  |  |  |  |  |
| South Atlantic | 0.42 | 0.59 | 1.02 | 3.62 | 0.43 |
| Atlanta | 0.44 | 0.55 | 0.99 | 4.39 | 0.54 |
| Miami | 0.46 | 0.89 | 1.35 | 7.07 | 0.57 |
| Washington, DC | 0.44 | 0.68 | 1.13 | 5.45 | 0.44 |
| East South Central | 0.52 | 0.48 | 1.00 | 2.48 | 0.36 |
| West South Central | 0.50 | 0.63 | 1.13 | 4.54 | 0.44 |
| Dallas | 0.33 | 0.58 | 0.92 | 4.13 | 0.58 |
| Houston | 0.40 | 0.73 | 1.12 | 6.52 | 0.41 |
| Midwest |  |  |  |  |  |
| East North Central | 0.45 | 0.57 | 1.01 | 4.05 | 0.42 |
| Chicago | 0.40 | 0.81 | 1.21 | 7.10 | 0.41 |
| Detroit | 0.35 | 0.52 | 0.87 | 1.54 | 0.61 |
| West North Central | 0.44 | 0.57 | 1.01 | 4.95 | 0.38 |
| West |  |  |  |  |  |
| Mountain | 0.37 | 0.43 | 0.80 | 4.53 | 0.40 |
| Denver | 0.40 | 0.38 | 0.78 | 2.64 | 0.62 |
| Pacific | 0.43 | 0.75 | 1.18 | 6.00 | 0.36 |
| Los Angeles | 0.46 | 0.70 | 1.16 | 4.48 | 0.46 |
| San Diego | 0.39 | 0.55 | 0.94 | 3.61 | 0.41 |
| San Francisco | 0.44 | 0.88 | 1.32 | 11.70 | 0.30 |
| Seattle | 0.43 | 0.69 | 1.12 | 5.56 | 0.31 |

PrEP: pre-exposure prophylaxis
\*Degree is the number of persistent male partners measured on the day of the survey completion. Total degree includes both main and casual persistent partners, main degree includes main partners only, and causal degree includes casual partners only.
†Regions include the nine US Census Bureau divisions but exclude the major listed cities within these divisions, where relevant. The New England division excludes Boston; the Middle Atlantic division excludes New York City and Philadelphia; the South Atlantic division excludes Atlanta, Miami, and Washington, DC; the East South Central division has no exclusions; the West South Central division excludes Dallas and Houston; the East North Central division excludes Chicago and Detroit; the West North Central division has no exclusions; the Mountain division excludes Denver; the Pacific division excludes Los Angeles, San Diego, San Francisco, and Seattle.

We found evidence of individual-level PrEP-related behavioral differences for multiple outcomes. Adjusting for demographics, individual PrEP use was associated with a higher total degree (adjusted IRR [aIRR]=1.70; 95% CrI, 1.57–1.83), casual degree (aIRR=2.34; 95% CrI, 2.14–2.56), and count of one-time partners (aIRR=4.62; 95% CrI, 3.94–5.44), and with a lower prevalence of consistent condom use in one-time partnerships (aIRR=0.60; 95% CrI, 0.53–0.69) (Table 4).

**Table 4.** Bivariable and Multivariable Associations of Individual PrEP Usage and Various Outcomes of HIV-negative ARTnet Participants

| Outcome | Unadjusted |  | Adjusted for Age and Race |  |
| --- | --- | --- | --- | --- |
|  | IRR (95% CrI) | % of Posterior >1.00 | IRR (95% CrI) | % of Posterior >1.00 |
| Total degree<sup>†</sup> | 1.73 (1.61, 1.86) | 100.00 | 1.70 (1.57, 1.83) | 100.00 |
| Main degree | 0.94 (0.82, 1.07) | 17.65 | 0.96 (0.84, 1.10) | 29.73 |
| Casual degree | 2.45 (2.24, 2.69) | 100.00 | 2.34 (2.14, 2.56) | 100.00 |
| Count of one-time partners<sup>‡</sup> | 4.62 (3.94, 5.40) | 100.00 | 4.62 (3.94, 5.44) | 100.00 |
| Always using condoms in one-time partnership(s) | 0.58 (0.51, 0.66) | 0.00 | 0.60 (0.53, 0.69) | 0.00 |

IRR: incidence rate ratio; CrI: credible interval; PrEP: pre-exposure prophylaxis
<sup>†</sup>Degree is the number of persistent male partners measured on the day of the survey completion. Total degree includes both main and casual persistent partners, main degree includes main partners only, and causal degree includes casual partners only.
<sup>‡</sup>Because of overdispersion, approximated using negative binomial distribution.

We also found evidence that community-level behavioral differences occurred independent of individual-level PrEP-related behavioral differences. Adjusting for demographics, community PrEP coverage was associated with higher total degree (aIRR=3.55; 95% CrI, 1.87– 7.54), casual degree (aIRR=6.99; 95% CrI, 3.04–19.36), and count of one-time partners (aIRR=34.42; 95% CrI, 9.98–144.11) (Table 5). Adjusting for demographics and individual PrEP use, the primary associations with community PrEP coverage were attenuated for total degree (aIRR=1.73; 95% CrI, 0.92–3.44), casual degree (aIRR=2.05; 95% CrI, 0.90–8.54), and count of one-time partners (aIRR=1.90, 95% CrI, 0.46–8.54); adjustment for individual PrEP use reduced these aIRRS by 52%, 71%, and 94%, respectively. However, important patterns still remained after adjustment for individual PrEP use: for the associations between community PrEP coverage and total degree, casual degree, and count of one-time partners, approximately 95%, 95%, and 82% of posterior distributions resulting from model simulations were above 1.00, signaling a positive relationship between community PrEP coverage and these outcomes. Community PrEP coverage was also related to consistent condom use (aIRR=1.68; 95% CrI, 0.86–3.35) in the fully adjusted model, though there was greater uncertainty.

**Table 5.**
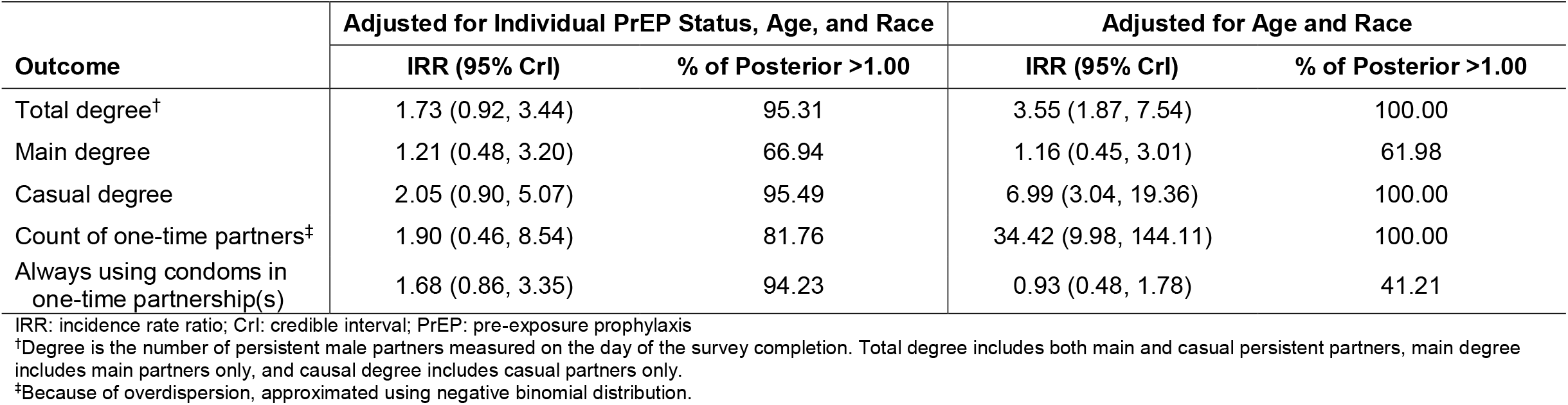
Multivariable Associations of Community-Level PrEP Usage and Various Outcomes of HIV-negative ARTnet Participants

## DISCUSSION

This study provides evidence of the correlation between community PrEP coverage and individual-level sexual behaviors among US MSM. We found that individual behaviors may be influenced by broader PrEP coverage at the community level independent of individual PrEP use. Individual PrEP use explained much of the observed differences in sexual behavior between individuals, but differences in sexual network degree, condom use, and one-time partnership formation by community PrEP coverage were observed even after adjusting for individual PrEP use. If this association is causal, it would support the hypothesis that MSM in communities with high PrEP coverage may have altered their behavior due to the scale-up of PrEP in their communities, independent of behavioral differences driven by their own PrEP use. This draws attention to the need for programs focused not only on increasing PrEP adherence and persistence among active PrEP users, but also on facilitating PrEP initiation for indicated MSM not currently using PrEP in cities with higher or increasing PrEP coverage.

Consistent with other studies,^9^ we found that individual PrEP use was linked to differences in sexual behavior. Specifically, we found that PrEP users had more one-time partners, used condoms less often, and had increased sexual network degree. These findings align with the theory that being on PrEP directly protects an HIV-negative individual from HIV acquisition, and this decrease in individual risk may drive changes to sexual behaviors (such as having more one-time partners). However, the directionality here may go both ways, because sexual behaviors such as condomless sex and multiple partners are indications for starting PrEP;^17^ the behaviors we observed in PrEP users in our analysis may have preceded PrEP initiation.

Our results additionally complement the few studies that have examined potential changes in sexual behavior among MSM not using PrEP but living in cities with high PrEP coverage.^12–14^ Our study expands on prior work by evaluating other sexual behaviors, including several sexual network outcomes. PrEP may be unequally distributed across a sexual network,^3^ and network attributes play a key role in HIV dynamics;^18^ network outcomes such as those we considered are crucial for evaluating the broader population impact of PrEP programs. This is a major strength of our study. Using multiple sexual behavior and network measures, we found evidence of differences in sexual behavior by community PrEP coverage. While most of the observed associations between community PrEP coverage and sexual behaviors were linked to individual PrEP use, notable associations remained after adjusting for individual PrEP use. This fits with the concept that for individuals not on PrEP, high PrEP coverage in their community may indirectly protect them from HIV acquisition, but this protection is weaker than the direct protection they would receive from PrEP.

Research examining both the direct and indirect effects of the scale-up of PrEP programs is important for developing effective disease prevention programs. While increasing PrEP coverage in a community can protect individuals either directly through PrEP use or indirectly by lower incidence (and ultimately prevalence) of HIV in their community, behavioral changes resulting from increased community-level PrEP coverage may mitigate some of these potential benefits. Increased rates of one-time partnerships, for example, can increase the risk of HIV acquisition among those not using PrEP. Further, protection against HIV is dependent on prevention-effective PrEP adherence. Individuals who discontinue PrEP (but do not alter their sexual behavior patterns) or have suboptimal adherence are not fully protected against HIV; in these cases, behavioral changes following PrEP initiation can increase the risk of HIV acquisition.

This study provides empirical evidence of potential community-level differences in behavior associated with PrEP coverage. Our findings suggest that MSM are making informed decisions regarding their behavioral risk and sexual wellness, and that these decisions may be affected by population-level factors. We do not interpret our findings as evidence that more prevention efforts should be made to reduce condomless sex in high PrEP coverage settings, but instead that PrEP use remains vital to reducing HIV transmission in communities. Our findings highlight that sexual behavior decisions do not occur only at an individual level, but also at a dyadic, network, and community level. Individual decisions about sexual behavior are not independent of partner behaviors; dyadic decisions impact PrEP eligibility, use, and resulting sexual behaviors.^19^ Though individuals may have agency over their sexual behavior, they remain affected by the decisions of the broader sexual network (e.g., the sexual behaviors of their partner’s partners). This has implications for PrEP delivery. Differences in sexual behavior related to PrEP use highlight the importance of eliminating PrEP barriers for all populations. For example, MSM who are indicated for but not currently on PrEP may be more likely to be Black,^20^ and Black MSM are at disproportionately high risk of contracting HIV.^1^ Notably, the increased HIV risk among Black MSM is not attributable to individual-level behaviors, but rather to network factors and socioeconomic and treatment disparities.^21^ Additionally, gaps in eligibility and initiation of PrEP among Black MSM are partly driven by provider racial bias, where clinicians may consider Black patients more likely than white patients to engage in condomless sex if prescribed PrEP, and therefore are less willing to provide PrEP to Black MSM.^22^ Our findings therefore underscore the need for public health programs focused on increasing PrEP initiation, adherence, and persistence, particularly in subpopulations that have been underserved by PrEP delivery to date.

Our findings also have implications for STI prevention. Beyond reducing HIV transmission, the prevention of STIs remains important: in addition to affecting overall sexual health, STIs can have adverse sequelae, increase the risk of HIV acquisition or transmission,^23^ and potentially divert resources away from HIV prevention and treatment. Substantial increases in STIs have been noted in the PrEP era; for example, rates of syphilis among US men increased by nearly 150% during 2015–2019.^24^ More frequent STI screening could alleviate the potential effects of PrEP-related differences in sexual behavior on STI incidence,^25^ but additional research, such as the impact of potential STI prophylaxis or vaccination on STI incidence in the context of community-level behavioral differences and high PrEP coverage, is needed.

Behavioral studies are necessary to understand the effects of PrEP use, but inferences drawn from these studies require consideration of how sexual behaviors and norms operate in a community. For example, while MSM not on PrEP who live in communities with high PrEP coverage may alter their sexual behaviors because they feel indirectly protected against HIV by men in their community being on PrEP, this assumes that MSM know that others in their sexual network/community are using PrEP. This may not be the case given the potential stigma associated with taking PrEP.^26^ Also, other factors may be operating at a community level beyond PrEP-related community-level sexual behaviors, such as culture around sexual risk, norms around HIV prevention, or differences in sexual networks by community. Community-level factors including, but not limited to, neighborhood gay presence,^27^ peer support for condom use,^28^ and social network norms and attitudes^29^ have been shown to drive HIV prevention behavior among MSM. Furthermore, differences in community sexual norms may have predated PrEP uptake and PrEP-era behavioral studies may not fully capture these long-term factors. Lastly, community-level viral suppression may be related to both PrEP use and community-level behaviors (e.g., communities with high PrEP coverage may also have high viral suppression). These issues should be considered when designing and interpreting studies examining community-level differences related to PrEP coverage.

### Limitations

First, the data used for this analysis are cross-sectional, with limited ability to establish causal relationships between community PrEP coverage and sexual behavior. Future studies should use longitudinal data to further examine the relationship between community PrEP coverage and behavioral outcomes (e.g., examine the changes in behavior of MSM in communities with PrEP coverage levels increasing over time). Second, our study used self-reported sexual behavior measures. There may be underreporting of behaviors, especially if study participants felt it was favorable to indicate use of HIV prevention behaviors. However, these data were collected via a web-based survey, reducing the possibility of social desirability bias.^30^ Also, while attitudes about HIV prevention behavior may differ between communities, it is unlikely that this would drive our findings. Third, our results are dependent on the accuracy of community PrEP coverage. To reduce the potential impact of this limitation on our findings, we estimated community PrEP coverage using a binomial model of individual PrEP use within a multilevel Bayesian model. This allowed us to retain the uncertainty of community PrEP coverage estimation in our models. Additionally, we found that the patterns of community-level PrEP coverage were comparable to NHBS estimates. Finally, ARTnet is not representative of all US MSM, especially of non-Hispanic Black MSM, who made up only 4.1% of our study sample. Future studies should focus on this key subpopulation.

## Conclusions

This is the first study to consider variable HIV PrEP coverage and sexual behaviors of US MSM using a variety of sexual behavior outcomes. Using multiple behavioral outcomes, we provided evidence for the hypothesis that sexual behavior may differ with variations in community PrEP coverage independent of individual PrEP use. Studies addressing this topic are increasingly relevant as PrEP use becomes more common. Ongoing assessment of community-level PrEP-related sexual behaviors is needed to guide public health recommendations, with supplemental HIV and STI prevention efforts focused on mitigating the potential effects of both individual- and community-level behavioral changes among PrEP users, their sexual partners not using PrEP, and the broader communities where PrEP coverage is high or increasing.

## Data Availability

Model code and data are available at the indicated git repository.

