## Supplementary Appendix for "Correlations Between Community-Level HIV Preexposure Prophylaxis Coverage and Individual-Level Sexual Behaviors among US Men Who Have Sex with Men"

### Supplementary Information: Statistical Analyses

This appendix contains information about the R packages used in this study. Statistical models were fit with the *rethinking* package,[1] which uses the STAN Markov Chain Monte Carlo Sampler to estimate model coefficients.[2]

### Supplementary Information: Measures

This appendix also contains information about ARTnet-derived estimates of current PrEP and National HIV Behavioral Surveillance System (NHBS)-derived estimates of city-level PrEP use. To validate the accuracy of the ARTnet-derived values of current PrEP use, we compared our PrEP estimates to 2017 NHBS estimates of city-level PrEP for 15 cities (NHBS data was only available for major cities and not for the nine census divisions).[3] Though NHBS PrEP coverage estimates were higher than ARTnet, the patterns between cities were similar: San Francisco had the highest current PrEP coverage in both NHBS and ARTnet, and Detroit had the lowest coverage according to NHBS and the second lowest coverage according to ARTnet (Supplemental Figure 1). Coverage estimates of were higher in NHBS in part because NHBS estimates represented only the proportion of MSM in 2017 who had a negative HIV test result at the time of the interview, did not report a previous HIV-positive test result, had either one male sex partner who was HIV-positive or multiple male sex partners in the past 12 months, and reported either condomless anal sex or a bacterial STI in the past 12 months, whereas ARTnet estimates were less restrictive, representing only HIV-negative and ever-tested MSM.

**Supplemental Figure 1**. Comparison of ARTnet-derived and National HIV Behavioral Surveillance System (NHBS)-derived values of city-level PrEP coverage for 15 major US cities. ARTnet values represent the proportion of HIV-negative and ever tested MSM currently on PrEP during 2017–2019. NHBS values represent the proportion of MSM in 2017 who are at risk for HIV infection (likely to meet clinical indications for PrEP, defined as men who had a negative HIV test result at the time of the interview, did not report a previous HIV-positive test result, had either one male sex partner who was HIV-positive or multiple male sex partners in the past 12 months, and reported either condomless anal sex or a sexually transmitted bacterial infection in the past 12 months) and reported using PrEP during 2017.

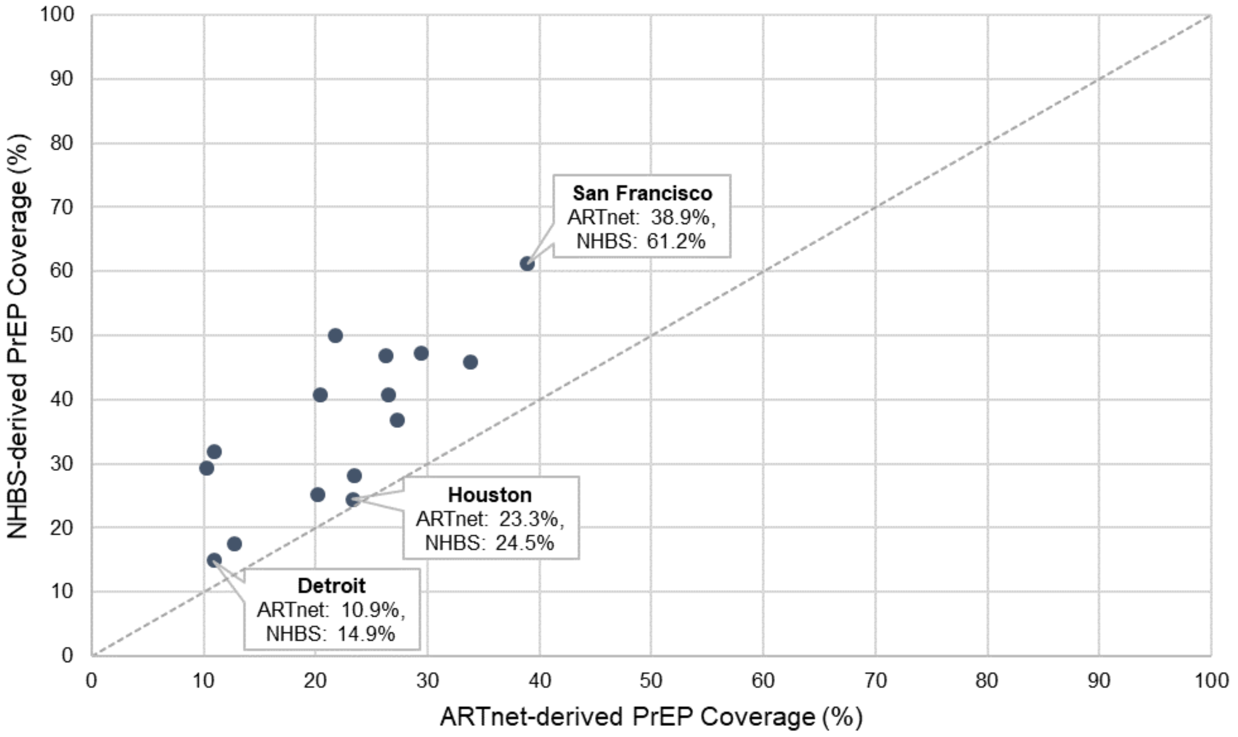

**Supplemental Table 1**. Sexual Network and Sexual Behavior Parameters for HIV-negative ARTnet Participants by Region/City, Stratified by Current PrEP Status

| | **Currently on PrEP (n=631)** | | | | | **Not on PrEP (Includes Non-Current and Never Used PrEP) (n=2,628)** | | | | |
|---|---|---|---|---|---|---|---|---|---|---|
| | **Average Main Degree*** | **Average Casual Degree** | **Average Total Degree** | **Average Count of One-Time Partnerships** | **Proportion of Individuals Who Always Used Condoms in One-Time Partnership** | **Average Main Degree** | **Average Casual Degree** | **Average Total Degree** | **Average Count of One-Time Partnerships** | **Proportion of Individuals Who Always Used Condoms in One-Time Partnership** |
| Total | 0.41 | 1.17 | 1.58 | 12.80 | 0.30 | 0.43 | 0.48 | 0.91 | 2.78 | 0.51 |
| Census Division/City† | | | | | | | | | | |
| Northeast | | | | | | | | | | |
| New England | 0.43 | 0.79 | 1.21 | 9.21 | 0.41 | 0.41 | 0.38 | 0.80 | 2.19 | 0.42 |
| Boston | 0.47 | 1.00 | 1.47 | 8.29 | 0.52 | 0.41 | 0.41 | 0.82 | 2.28 | 0.58 |
| Middle Atlantic | 0.32 | 1.11 | 1.43 | 13.00 | 0.51 | 0.47 | 0.47 | 0.94 | 3.08 | 0.51 |
| New York City | 0.27 | 1.27 | 1.54 | 12.60 | 0.34 | 0.43 | 0.46 | 0.88 | 2.74 | 0.70 |
| Philadelphia | 0.13 | 0.50 | 0.63 | 6.25 | 0.31 | 0.46 | 0.51 | 0.97 | 1.76 | 0.52 |
| South | | | | | | | | | | |
| South Atlantic | 0.42 | 1.09 | 1.51 | 13.20 | 0.26 | 0.42 | 0.53 | 0.95 | 2.29 | 0.48 |
| Atlanta | 0.41 | 0.91 | 1.31 | 11.80 | 0.48 | 0.45 | 0.44 | 0.89 | 2.12 | 0.58 |
| Miami | 0.29 | 1.71 | 2.00 | 13.30 | 0.29 | 0.48 | 0.77 | 1.25 | 6.17 | 0.61 |
| Washington, DC | 0.49 | 1.09 | 1.57 | 10.80 | 0.26 | 0.43 | 0.54 | 0.97 | 3.54 | 0.56 |
| East South Central | 0.53 | 0.90 | 1.42 | 5.42 | 0.14 | 0.52 | 0.41 | 0.93 | 1.98 | 0.43 |
| West South Central | 0.44 | 1.00 | 1.44 | 14.20 | 0.33 | 0.51 | 0.55 | 1.06 | 2.65 | 0.49 |
| Dallas | 0.65 | 1.06 | 1.71 | 8.06 | 0.63 | 0.25 | 0.46 | 0.72 | 3.13 | 0.56 |
| Houston | 0.35 | 1.47 | 1.82 | 12.50 | 0.13 | 0.41 | 0.50 | 0.91 | 4.71 | 0.58 |
| Midwest | | | | | | | | | | |
| East North Central | 0.32 | 0.94 | 1.26 | 9.83 | 0.28 | 0.47 | 0.49 | 0.97 | 2.90 | 0.47 |
| Chicago | 0.48 | 1.60 | 2.08 | 16.20 | 0.26 | 0.38 | 0.48 | 0.85 | 3.30 | 0.52 |
| Detroit | 0.60 | 1.40 | 2.00 | 4.00 | 0.75 | 0.32 | 0.42 | 0.73 | 1.24 | 0.59 |
| West North Central | 0.50 | 0.94 | 1.44 | 11.10 | 0.15 | 0.43 | 0.49 | 0.92 | 3.68 | 0.46 |
| West | | | | | | | | | | |
| Mountain | 0.17 | 1.17 | 1.33 | 11.60 | 0.21 | 0.39 | 0.35 | 0.74 | 3.69 | 0.44 |
| Denver | 0.50 | 1.00 | 1.50 | 6.83 | 0.50 | 0.39 | 0.31 | 0.69 | 2.12 | 0.64 |
| Pacific | 0.46 | 1.27 | 1.73 | 18.40 | 0.21 | 0.42 | 0.60 | 1.02 | 2.40 | 0.45 |
| Los Angeles | 0.36 | 1.31 | 1.67 | 11.20 | 0.34 | 0.50 | 0.47 | 0.97 | 1.96 | 0.53 |
| San Diego | 0.40 | 0.90 | 1.30 | 10.10 | 0.31 | 0.39 | 0.46 | 0.85 | 1.95 | 0.46 |
| San Francisco | 0.46 | 1.63 | 2.09 | 27.30 | 0.10 | 0.44 | 0.40 | 0.84 | 1.69 | 0.58 |
| Seattle | 0.46 | 1.38 | 1.85 | 11.00 | 0.25 | 0.41 | 0.33 | 0.75 | 2.80 | 0.35 |

PrEP: pre-exposure prophylaxis
*Degree is the number of persistent male partners measured on the day of the survey completion. Total degree includes both main and casual persistent partners, main degree includes main partners only, and causal degree includes casual partners only.
†Regions include the nine US Census Bureau divisions but exclude the major listed cities within these divisions, where relevant. The New England division excludes Boston; the Middle Atlantic division excludes New York City and Philadelphia; the South Atlantic division excludes Atlanta, Miami, and Washington, DC; the East South Central division has no exclusions; the West South Central division excludes Dallas and Houston; the East North Central division excludes Chicago and Detroit; the West North Central division has no exclusions; the Mountain division excludes Denver; the Pacific division excludes Los Angeles, San Diego, San Francisco, and Seattle.
